# Trends in voice characteristics in patients with heart failure (VENTURE) in Switzerland: Protocol for a longitudinal observational pilot study

**DOI:** 10.1101/2023.03.06.23286682

**Authors:** Fan Wu, Matthias Nägele, David Cleres, Thomas Haider, Elgar Fleisch, Frank Ruschitzka, Andreas Flammer, Filipe Barata

## Abstract

**Introduction:** Heart Failure (HF) is a major health and economic issue worldwide. HF-related expenses are largely driven by hospital admissions and re-admissions, many of which are potentially preventable. Current self-management programs, however, have failed to reduce hospital admissions. This may be explained by their low predictive power for decompensation and high adherence requirements. Slight alterations in the voice profile may allow to detect decompensation in HF patients at an earlier stage and reduce hospitalizations. This pilot study investigates the potential of voice as a digital biomarker to predict health status deterioration in HF patients.

**Methods and analysis:** In a two-month longitudinal observational study, we collect voice samples and HF-related quality-of-life questionnaires from 35 stable HF patients. Patients use our developed study application installed on a tablet at home during the study period. From the collected data, we use signal processing to extract voice characteristics from the audio samples and associate them with the answers to the questionnaire data. The primary outcome will be the correlation between voice characteristics and HF-related quality-of-life health status.

**Ethics and dissemination:** The study was reviewed and approved by the Cantonal Ethics Committee Zurich (BASEC ID:2022-00912). Results will be published in medical and technical peer-reviewed journals.

## 1 Introduction

Heart Failure (HF) is characterized by the heart’s incapacity to pump sufficient blood to meet the body’s metabolic needs [1]. It afflicts over 64.3 million people worldwide and is increasing in prevalence [2]. According to data from 2015 to 2018, an estimated 6 million American adults suffered from HF [3]. High hospital readmission rates not only pose a tremendous burden on a patient’s health status, morbidity, and mortality but also significantly increase national healthcare costs [4]. The average cost per patient per hospitalization in Europe is approximately €10,000 [5]. The total cost of care for heart failure in 2020 is estimated at $43.6 billion in the US [6]. While HF hospitalizations constitute 60% of the total expenditures associated with HF [1], many hospital admissions are considered preventable [7]. It implies that these costs, mainly caused by repeated and lengthy hospitalizations, can be significantly reduced.

To handle this amount of patients and to reduce costs, self-management programs empower patients as they can actively contribute to managing their disease and participate in continuous education, self-care promotion, and therapy adherence [8]. Current self-management programs advise patients to monitor their daily weight and call their physician if they experience rapid weight gain or clinical signs of congestion such as peripheral edema [9]. Self-management programs (e.g., weight, blood pressure), however, have limited predictive power for decompensation and require high adherence and constant commitment [10, 11]. Not surprisingly, clinical trials involving a broader population and using traditional self-management programs have not yet reduced hospitalizations [12].

On this basis, we argue that passive, non-invasive, and user-friendly tools for HF patients are required to facilitate self-management. Remarkably, the monitoring of voice has the potential to identify physiological changes and health conditions [13]. Speech sounds are created by pressure in the lungs and then propagated through the vocal tract [14]. As decompensation approaches, i.e., when the heart can no longer maintain efficient circulation, HF patients develop fluid retention throughout the body, including the lungs. In particular, the vocal folds consist of thin tissue layers that might be particularly sensitive to HF-related edema [15]. We hypothesize that the vocal folds are more sensitive to fluid accumulation than weight changes. Thus, voice monitoring may allow timely detection of fluid volume imbalance (e.g., volume overload) and congestion and prevent acute decompensated heart failure events that require hospitalization.

Mobile devices such as smartphones and tablet computers have become ubiquitous in our lives, e.g., 85% of US adults owned a smartphone in 2021 [16]. Additionally, voice-based conversational agents (VCAs), such as Apple’s Siri, are now used on more than 2.5 billion smartphones, tablets, smart speakers, and wearable devices. To this end, voice samples of HF patients can be easily collected at home from their voice commands to VCAs. Thanks to the widespread use of information and communication technologies, voice analysis can be used as a scalable, tailored, and cost-effective health monitoring system. Further, a voice monitoring system may ease the interaction with technology for the elderly, as studies have shown the use of voice interfaces appears to be easier and more acceptable for older and frail people [17–19]. Moreover, the voice monitoring system can especially support individuals with low literacy or intellectual [20], and motor or cognitive disabilities [21].

This research aims to contribute to developing a digital vocal monitoring tool as a new biomarker for HF. We use voice features collected from mobile devices and integrate voice analysis into patient self-care. Voice can be readily captured as a health parameter to identify the likelihood of progression of HF. Collecting voice samples passively while being of clinically valuable quality can contribute to two major issues of remote care for HF patients: the lack of adherence and the need for robust and clinically relevant measurements in patients’ homes.

### 1.1 Hypothesis and objectives

We hypothesize that fluctuations in voice features extracted from recorded voice samples from HF patients using smart devices can discriminate HF-related health status changes. The primary objective is to investigate how the variations in the overall health status of HF patients correlate with the fluctuations in voice characteristics.

The secondary objectives of this study are to:

- compare voice characteristics with the current standard of care (i.e., weight and blood pressure).
- investigate how the health status correlates with the combination of nocturnal cough frequencies and physiological data (e.g., steps).
- investigate how voice characteristics fluctuate with mental health conditions (e.g., depression and anxiety).
- evaluate the technology acceptance of the developed application in HF patients.

## 2 Methods

### 2.1 Study design

In a 2-month prospective, longitudinal observational study, we collect voice samples daily and calculate voice characteristics fluctuations from those. The primary endpoint is the potential association between voice characteristics and health status variations. The variations in health status are assessed using the 23-item Kansas City Cardiomyopathy Questionnaire (KCCQ) [22]. The KCCQ quantifies physical limitations, symptoms, self-efficacy, social interference, and quality of life of HF patients. While the KCCQ is valid for two weeks, this study includes the HF Symptom Tracker (HFaST) questionnaire to assess daily symptoms in HF patients [23]. The study duration per patient is 57 days, respectively 56 nights, which enables five repeated measurements of the KCCQ score.

To compare the new vocal biomarker to current surrogate biomarkers (i.e., weight and blood pressure), HF patients measure weight and blood pressure daily following Swiss Heart Foundation self-management guidelines [24]. Besides, we collect nocturnal cough frequency, physiological data, mental health, and technology acceptance as secondary research endpoints. Nocturnal cough monitoring data are collected every night with an application developed in our previous work [25–27]. The application has a cough detection model developed based on 26,166 cough samples from 94 adults with asthma. The model yielded an accuracy of 99.8% under 15 new patients. Physiological data (e.g., heart rate, steps, stress) are measured daily with a smartwatch, Garmin Vivoactive 4s. The mental health conditions (e.g., depression and anxiety) are assessed biweekly with the 9-item Patient Health Questionnaire (PHQ) [28] and the 7-item Generalized Anxiety Disorder (GAD) questionnaire [29]. Both questionnaires are brief, well-validated measures for detecting and monitoring depression and anxiety [30]. All the measurements are recorded using a Lenovo K10 tablet at home. At last, we apply the technology acceptance model [31] to examine whether our technology is acceptable and user-friendly.

### 2.2 Eligibility criteria

A selected group of HF patients will be studied to maximize the likelihood of health fluctuations observed in the biweekly KCCQ. Table 1 demonstrates the applied inclusion and exclusion criteria.

**Table 1.**
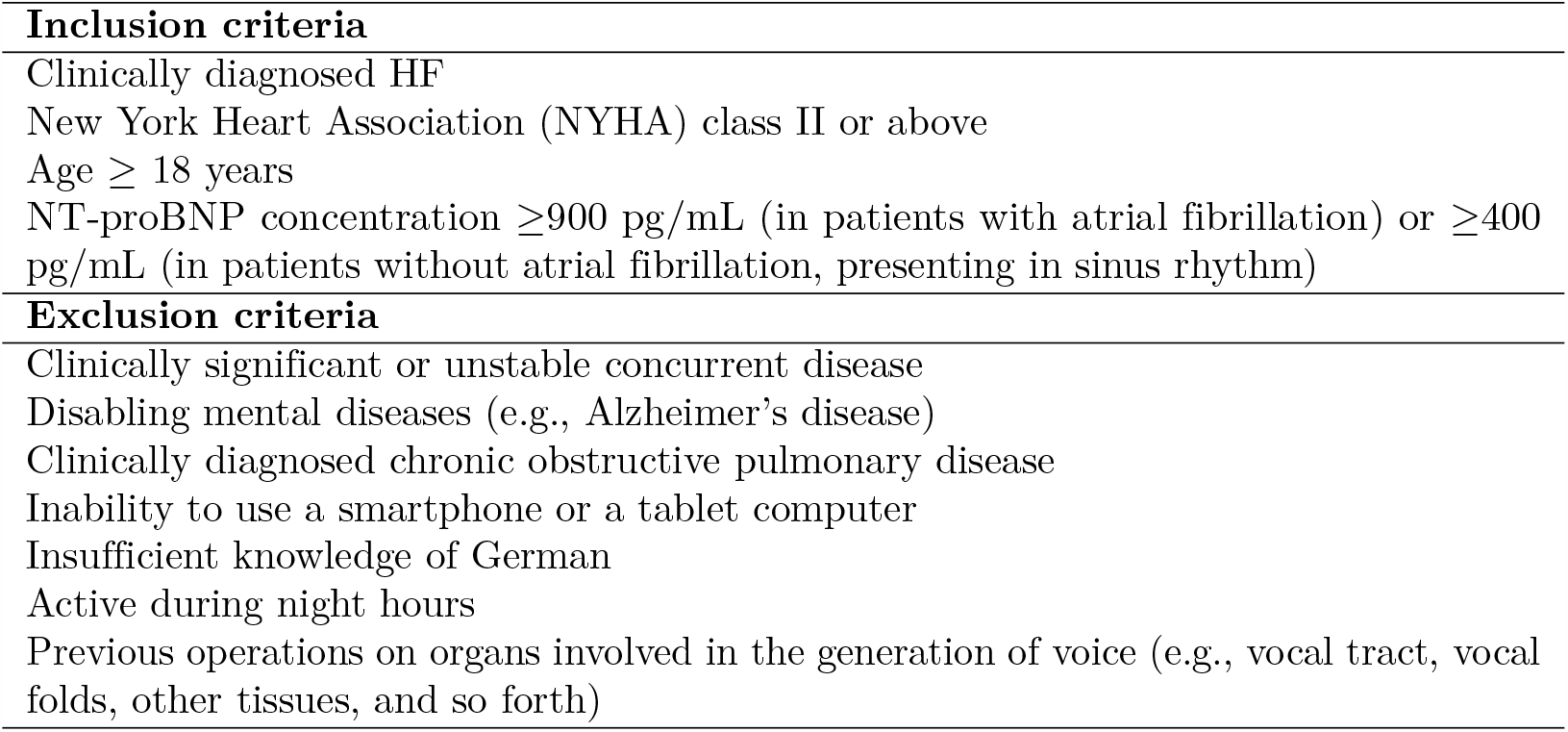
Inclusion and exclusion criteria Inclusion criteria.

### 2.3 Sample size

The study is powered on the primary objective: the association between the patient’s health status changes and daily collected voice feature fluctuations. We simulate the number of required patients to reach power (1-*β*) of 95% with a two-tailed type 1-error probability (*α*) of 5%, rejecting a zero bivariate correlation with a dropout rate of 15%. The R simulations (n=1000) relied on the MASS:mvrnorm function to generate two correlated random samples to fit a linear mixed-effects model using the lmerTest:lmer function [32, 33]. Based on previous studies [15, 34, 35], it is reasonable to assume: (i) a correlation of 0.3 between the voice characteristics and the KCCQ score, (ii) a mean KCCQ value in stable HF NYHA class II of 73±19, and (iii) a mean voice biomarker value of 141 ± 16 with five repeated KCCQ measures. The sample size is calculated at 35 patients, which gives a power of 95%. Assuming a dropout rate of 15%, we will recruit 41 patients.

### 2.4 Study setting & initial meeting

Patients are recruited at the University Hospital of Zurich (USZ) based on their clinical HF diagnosis and are included in the study. Health professionals perform medical assessments during baseline and follow-up visits. Table 2 shows the patient’s assessments and measurements. During the baseline visit, the study team will recruit patients with high concentrations of NT-proBNP. The study team will give informed consent to the eligible patients and ask for their willingness to participate in the study.

**Table 2.** Assessments and measurements through the study period.

| Phase | Study date | Key assessments / measurements |
| --- | --- | --- |
| Baseline visit | d0 | Medical assessments<br>Participant informed consent |
| Technical visit | d1 | Technical introduction & training<br>First data collection |
| Data collection | d2-d57 | Voice, weight, blood pressure, questionnaire,<br>nocturnal cough |
| Follow-up visit | d58 | Medical assessments<br>Blood draw<br>Technology acceptance questionnaire |

The first participant was included in September 2022. At the time of manuscript submission, two participants have completed the study. Data collection is expected to conclude in the third quarter of 2023. During the data collection phase, one patient is required to complete measurements at least 80% of the time throughout the study period (i.e., more than 45 days in 57 days). The KCCQ questionnaire should appear five times in one study phase, and the patient needs to finish it at least four times.

Medical assessments during baseline and follow-up visits include patient history (e.g., concomitant diseases), clinical examination (e.g., NYHA-class, NT-proBNP), and medical examination. The blood draw in the follow-up visit consists of sodium, potassium, creatinine, NT-proBNP, and a hematogram. The technical visit is planned to occur within 14 days after the baseline visit. The follow-up visit is planned within 14 days after the final data collection.

## 3 Data collection and management

### 3.1 Measures

During the study period, all measurements are taken using the tablet on which the study applications are installed. The study consists of active and passive measurements. The duration and frequency of the different measurements are summarized in Table 3.

**Table 3.** Duration of the daily and weekly, active and passive measurements.

| Measurement | Duration | Type | Frequency |
| --- | --- | --- | --- |
| Weight | 1 minute | active | daily |
| Voice - Vowels (/a/, /i/, /o/) | 1 minute | active | daily |
| Voice - 5 voice commands | 1 minute | active | daily |
| Voice - Nordwind passage | 2 minute | active | daily |
| Voice - Running speech | 20 seconds | active | daily |
| HFaST Questionnaire | 1 minute | active | daily |
| Control questions | 1 minute | active | daily |
| Blood Pressure | 2 minutes | active | daily |
| KCCQ Questionnaire | 3 minutes | active | biweekly |
| PHQ & GAD Questionnaire | 3 minutes | active | biweekly |
| Cough detection | the whole night | passive | daily |
| Physiological data (optional) | the whole day | passive | daily |

Each vowel is pronounced three times for five seconds. Voice commands are those used in the interaction with a VCA [36]. Control questions are designed to reduce the impact of confounding variables, i.e., noise when performing voice exercises, the tablet’s position during the night, the presence of an upper respiratory infection, and so forth. Physiological data is measured only when the participant agrees to wear the smartwatch.

#### Active measurements

The patients are required to measure their weight, perform voice exercises, fill out questionnaires, and measure their blood pressure in sequence. It takes around 10 minutes per day to perform all the measurements. Audios are recorded with a sampling rate of 44.1kHZ, 16 bits per sample, and the pulse-code modulation codec. Besides the routine daily measurements, the KCCQ, GAD, and PHQ questionnaires are alternately displayed in the study app, depending on the week. The KCCQ questionnaire appears on the 1st, 15th, 29th, 43rd, and 57th day, representing the health status in the last two weeks, while the questionnaires about depression and anxiety, GAD and PHQ, appear on the 8th, 22nd, 36th, and 50th day. The answers to all the questionnaires are consistently and uniformly assessed.

#### Passive measurements

Nocturnal cough frequency is measured automatically at night during patients’ sleeping time. Patients are instructed to place the tablet on the nightstand or near their bed. Optionally, patients are equipped with a smartwatch that passively collects continuous physiological data. The data from the smartwatch is automatically synchronized when the smartwatch is close enough to the study tablet.

### 3.2 Outcomes

The **primary outcome** is the association between voice characteristics and health status fluctuations in HF patients. We hypothesize that the voice contains sufficient correlates of health-related information and could serve as an effective monitoring biomarker. The **secondary outcomes** are:

- comparison of health status prediction using voice characteristics and current standard of care (i.e., weight and blood pressure).
- the correlation between health status fluctuations and the combination of nocturnal cough frequencies and physiological data.
- the correlation between voice characteristics and mental health status.
- the technology acceptance of the voice capture application.

### 3.3 Data management

We have designed a study application to collect all active measurements (e.g., audio data, blood pressure, weight, and questionnaire answers). Firstly, we store all measurements in a local SQLite database on Android. Then we transmit the measurements with the tablet’s metadata over a secure protocol (i.e., HTTPS) and save them remotely on a password-protected MySQL server hosted by ETH Zurich.

Nocturnal cough audio data is saved on the tablet, and only cough frequencies are uploaded to the server. The physiological data is measured with a smartwatch and uploaded to Labfront, which were utilized in psychological studies and clinical trials [37]. Participants’ smartwatches directly connect to the Labfront app via a Bluetooth connection. The study team will manually transfer data from the Labfront cloud to the database. Fig 1 demonstrates the data pipeline from the study app to the web server. Once a patient has completed the study, data are temporally saved on an encrypted local hard drive and then uploaded to ETH Zurich’s Leonhard Med Secure Scientific Platform [38] for data storage and analysis.

**Fig 1.**
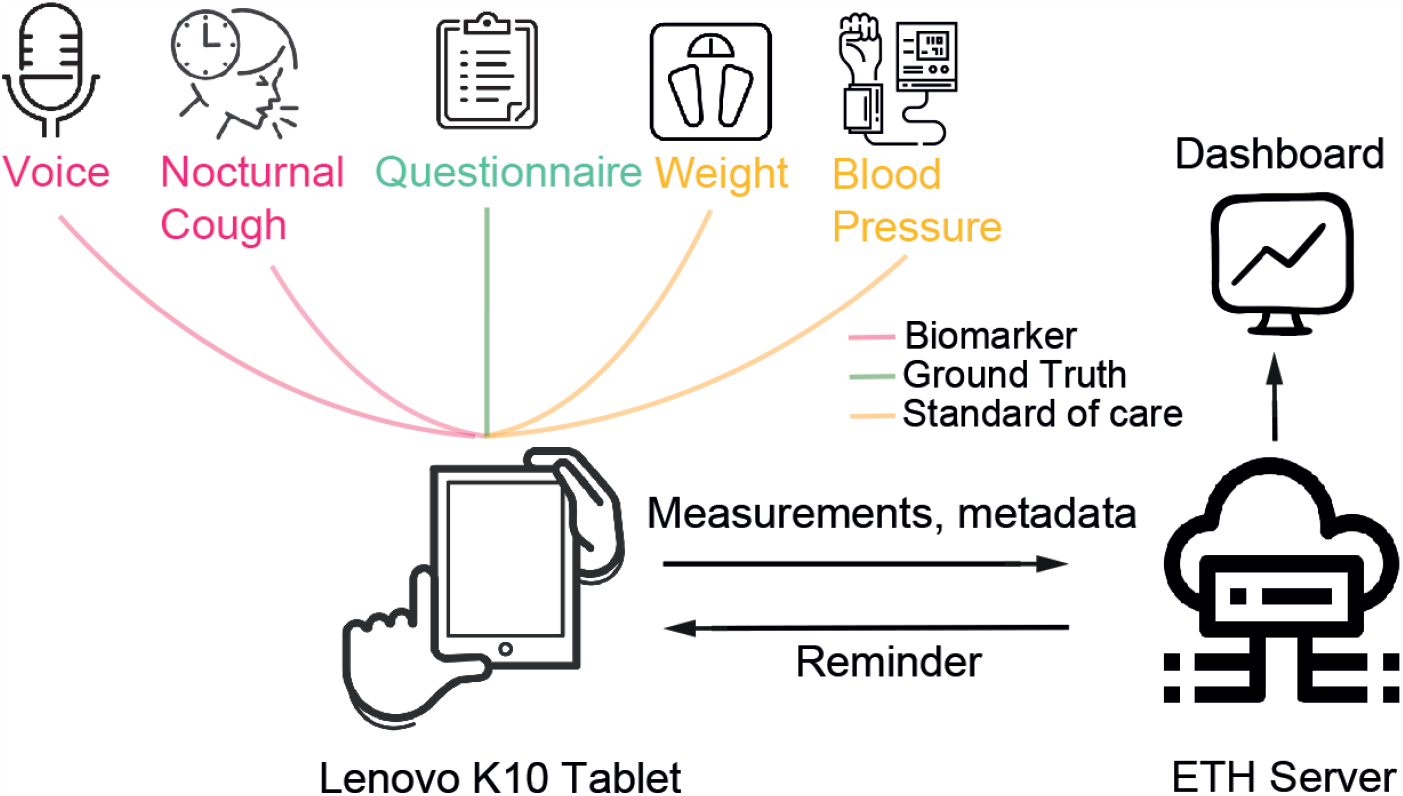
Data pipeline. Patients collect data every morning using the developed application on the Lenovo K10 tablet. All active measurements and nocturnal cough frequency are collected and transmitted to the ETH web server. The server integrates new data into the cloud and schedules reminders. Based on the data on the server, the dashboard visualizes and monitors measurements and metadata.

We have developed a dashboard for monitoring data and the status of patients. Physicians have access to the measured data of each patient (pseudonymized by a unique hash code) through a website. Fig 2 depicts the dashboard that visualizes measurements, tablet’s metadata, and study progress (e.g., 50% means the patient has conducted the study for one month).

**Fig 2.**
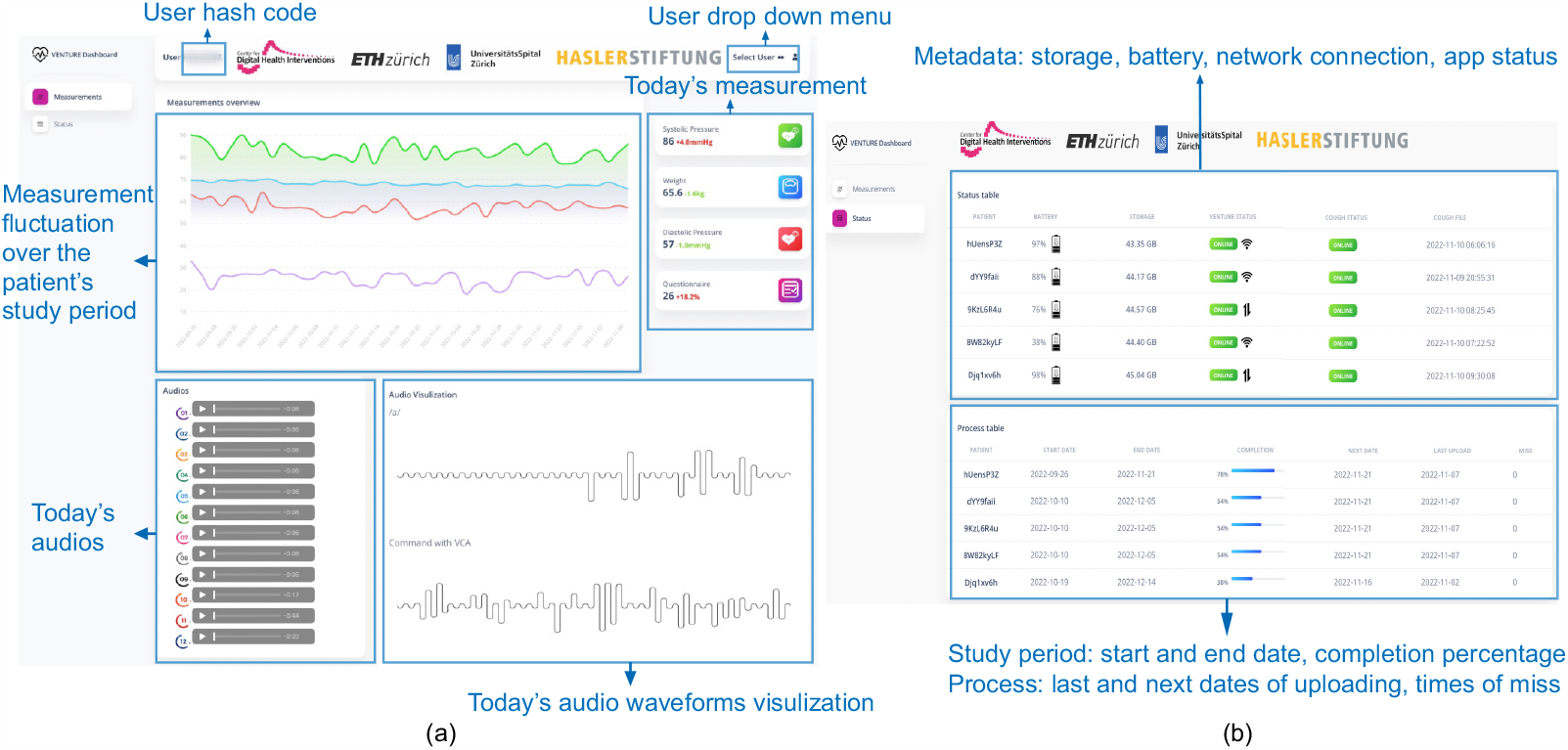
Dashboard. (a) The “Measurement” dashboard depicts today’s measurements and audio, numerical measurement fluctuations during the study period, and audio waveform visualization. (b) The “Status” dashboard shows the tablet’s metadata, e.g., storage, network connection, and the study progress.

All clinical data are taken from the electronic patient file and transferred to paper case report forms. Paper case report forms are stored in a locked room in the University Heart Center Zurich study center.

### 3.4 Retention

We promote adherence to the data collection phase using several methods. First, participants receive an artistic picture as a small reward after completing their daily measurements. The app changes pictures daily and depicts one scenic location worldwide with a description. Fig 3 shows four screenshots of the study application with English translation for readability. Texts in the application are originally written in German.

**Fig 3.**
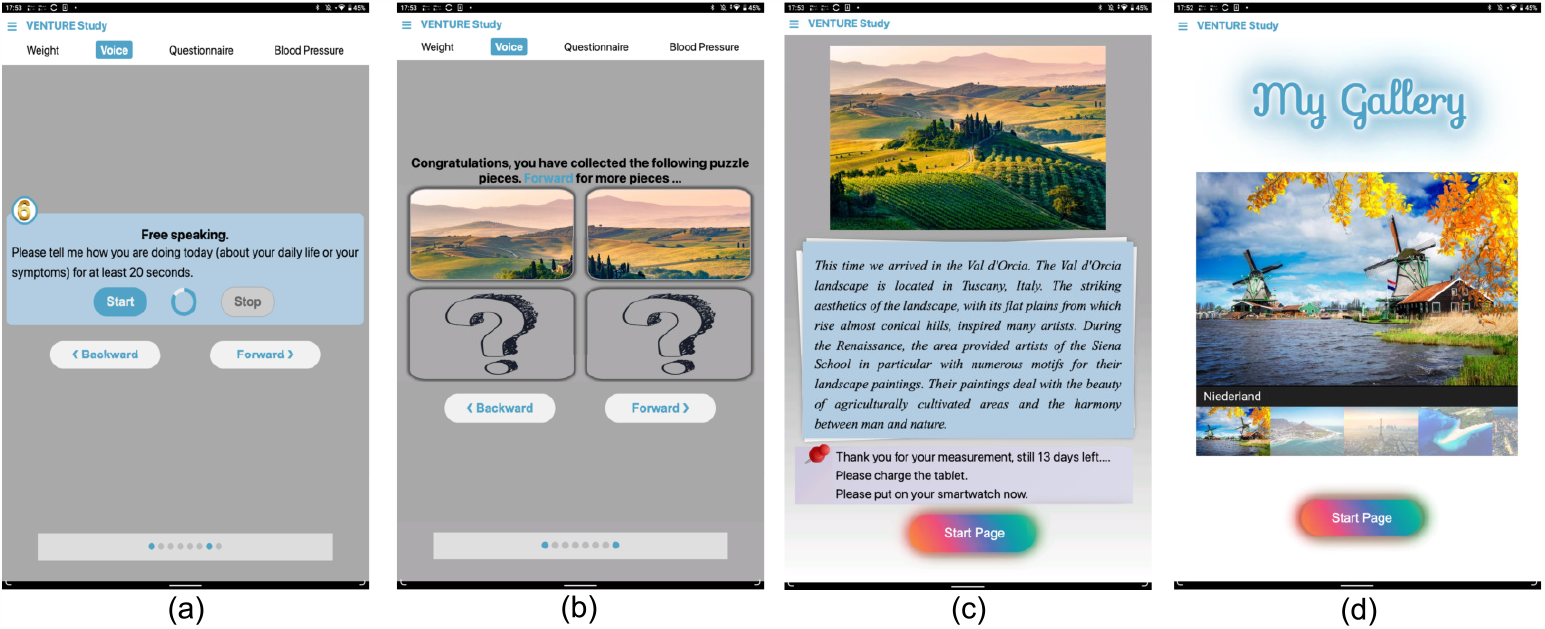
Four screenshots of the study application. (a) A screenshot of the voice recording procedure. (b) The participant obtains two pieces of the image after finishing two measurement sections. (c) After completing the measurement, the participant obtains a new image and its description as a reward. (d) The participant can view all collected images in the gallery.

Second, the study application involves a reminder system for measurement. If an unexpected issue happens, e.g., the participant fails to upload the data, the tablet has a low battery (≤ 40%), or there is no network connection, the participant will receive an SMS notification on their private phone or an email. In the event of non-adherence for two consecutive days, the study team will call the participant. If the tablet encounters an issue (e.g., it is turned off unintentionally by the patient), the study team will access the tablet with remote control and fix the issue.

Third, if the patient does not complete the biweekly KCCQ questionnaire on the given day, the app will display the questionnaire over the next three days until the patient finishes that. It maximizes the chance that the patient will complete the KCCQ questionnaire. If the patient has not completed the questionnaire after three days, it is considered a failed attempt, and the patient will be excluded from the study.

## 4 Data analysis

### 4.1 Feature Extraction

First, we use signal processing to calculate parameters that describe energy changes and amplitude changes in time and frequency from collected audio samples. Based on previous studies [39–41]) and pre-established libraries (i.e., Praat, librosa, and openSMILE), we will extract the following voice features, e.g., fundamental frequency, formants, jitter, shimmer [42], creak, and Mel-Frequency Cepstral Coefficients [43].

### 4.2 Correlation Analysis

We will investigate the correlation between voice characteristics and health status, represented by KCCQ scores, at between- and within-subject levels. We will perform the repeated measures correlation (rmcorr) method using a statistical programming package available in R (rmcorr) or Python (pingouin.r corr) [44].

### 4.3 Predictive Modeling

KCCQ scores constitute the ground truth, and the selected voice features serve algorithm features. We will predict the health status using typical classification methods, e.g., random forests, support vector machines, neural networks [45]. Afterwards, to further analyze and predict longitudinal time series data, we will explore recurrent neural networks and long short-term memory [46, 47].

### 4.4 Missing data

Various techniques for imputing missing data points will be considered (e.g., imputing mean, median, most frequent, or constant values) [48]. Depending on the type of variables, we will consider more advanced techniques (e.g., regression, interpolation, extrapolation, k-NN, multivariate imputation, and datawig) [49, 50].

## 5 Ethics considerations

The study was approved by Swissethics (BASEC-ID: 2022-00912). This research project will be conducted following the Declaration of Helsinki [51], the principles of Good Clinical Practice, the Human Research Act (HRA), the Human Research Ordinance (HRO) [52] as well as other locally relevant regulations.

## 6 Discussion

### 6.1 Novelty

Current studies have shown research potential for voice analysis in HF patients. Murton et al. analyzed the voice of HF patients treated for decompensated HF and showed changes in patient voice between baseline and study end measurements [15]. Sara et al. have shown that in HF patients, there is an association between invasively measured indices of pulmonary hypertension and a linear combination of the 223 acoustic features (e.g., Mel cepstrum representation and formant measures) extracted with “Vocalis Health” application [53]. Using the same application to extract voice features, Maor et al. have shown that vocal biomarkers calculated from patients’ recorded speech correlate with the likelihood of hospitalization and death [54]. Amir et al. compared voice recordings of 40 patients with acute decompensated HF to baseline recordings and successfully identified them as significantly different in 87.5% of cases [55].

While preliminary evidence suggests an association between voice signals and adverse outcomes in HF patients, previous studies have focused on the extreme states (i.e., decompensation and non-decompensation) but not on the continuous voice changes over time. Moreover, there is no evidence that voice characteristics would add value to the standard of care. It is crucial not only to assess whether the features of the voice are informative but also whether this measurement is complementary to the established measures. Very few researchers focused on the prediction power of voice, and there was no remote monitoring system based on a vocal biomarker.

Our approach differs from previous research in the following aspects. First, we look into how longitudinal changes in voice can be associated with changes in health status over time. Second, we assess the added value of voice monitoring compared to the current standard of care, weight, and blood pressure measurements. Third, we use advanced machine learning methods on real-world audio data to predict health deterioration.

### 6.2 Limitations

The study has certain limitations. First of all, voice analysis can be subject to various biases. Voice depends on different factors, among others, age, sex, and ethnicity. Since 70-80% of HF patients are male, the gender ratio is not set at 1:1 for better recruitment. Selection bias due to the technology adoption rate among seniors and German-speaking ability might also undermine generalizability. Besides, voice differs when the patient has upper respiratory infection symptoms in the study period. Thus, we propose the control question to control the confounding effects that might affect the voice.

Secondly, scales and blood pressure monitors are not uniformly provided. Patients use their own devices instead (unless they do not have them).

Thirdly, the applied cough detection model has yet to be developed based on data from HF patients and has not been used on tablets. To guarantee the functionality of the cough detection model, the tablet also records sound during the night. In this way, we can correct for wrongly counted coughs and post-hoc, acoustically verify the cough frequencies reported by our models to evaluate the performance of our models.

## 7 Conclusion

The proposed longitudinal study focuses on the association between continuous voice fluctuations and the health status of HF patients over time. We evaluate the potential of using voice characteristics to predict health deterioration. Furthermore, the comparison of voice analysis to the standard of care and patient feedback will reveal to what extent voice is accepted as a new monitoring tool. The study aims to lay the groundwork for using the voice to recognize clinical deterioration and initiate early intervention and management.

## Data Availability

Data cannot be shared publicly because of ethical requirements. Data are available from the Kantonale Ethikkommission (Cantonal Ethics Committee Zurich) BASEC ID:2022-00912 (contact via) for researchers who meet the criteria for access to confidential data.

## Author Contributions

**Conceptualization:** Elgar Fleisch, Andreas Flammer, Filipe Barata

**Data curation:** Fan Wu, Matthias Nägele

**Funding acquisition:** Filipe Barata

**Investigation:** Fan Wu, Matthias Nägele

**Methodology:** Fan Wu, Matthias Nägele, David Cleres, Thomas Haider, Elgar Fleisch, Andreas Flammer, Filipe Barata

**Project administration:** Elgar Fleisch, Andreas Flammer, Filipe Barata

**Resources:** Matthias Nägele, Thomas Haider, Andreas Flammer

**Software:** Fan Wu

**Supervision:** Elgar Fleisch, Andreas Flammer, Frank Ruschitzka, Filipe Barata

**Visualization:** Fan Wu

**Writing - original draft:** Fan Wu

**Writing - reviewing & editing:** Matthias Nägele, David Cleres, Thomas Haider, Elgar Fleisch, Andreas Flammer, Filipe Barata

## Acknowledgments

We would like to thank ETH Zurich’s statistical consulting service for their support on power calculation.

## References

1. Mangini S, Pires PV, Braga FGM, Bacal F. Decompensated heart failure. Einstein (Sao Paulo). 2013;11:383–391.

2. James SL, Abate D, Abate KH, Abay SM, Abbafati C, Abbasi N, et al. Global, regional, and national incidence, prevalence, and years lived with disability for 354 diseases and injuries for 195 countries and territories, 1990–2017: a systematic analysis for the Global Burden of Disease Study 2017. The Lancet. 2018;392(10159):1789–1858.

3. Virani SS, Alonso A, Aparicio HJ, Benjamin EJ, Bittencourt MS, Callaway CW, et al. Heart disease and stroke statistics—2021 update: a report from the American Heart Association. Circulation. 2021;143(8):e254–e743.

4. Urbich M, Globe G, Pantiri K, Heisen M, Bennison C, Wirtz HS, et al. A systematic review of medical costs associated with heart failure in the USA (2014–2020). Pharmacoeconomics. 2020;38(11):1219–1236.

5. Bundkirchen A, Schwinger RHG. Epidemiology and economic burden of chronic heart failure. European Heart Journal Supplements. 2004;6:D57–D60. doi:10.1016/J.EHJSUP.2004.05.015.

6. Heidenreich PA, Albert NM, Allen LA, Bluemke DA, Butler J, Fonarow GC, et al. Forecasting the Impact of Heart Failure in the United States. Circulation: Heart Failure. 2013;6:606–619. doi:10.1161/HHF.0B013E318291329A.

7. Michalsen A, König G, Thimme W. Preventable causative factors leading to hospital admission with decompensated heart failure. Heart. 1998;80:437–441. doi:10.1136/HRT.80.5.437.

8. Evangelista LS, Shinnick MA. What do we know about adherence and self-care? The Journal of cardiovascular nursing. 2008;23(3):250.

9. Gomberg-Maitland M, Baran DA, Fuster V. Treatment of Congestive Heart Failure: Guidelines for the Primary Care Physician and the Heart Failure Specialist. Archives of Internal Medicine. 2001;161:342–352. doi:10.1001/ARCHINTE.161.3.342.

10. Koulaouzidis G, Iakovidis DK, Clark AL. Telemonitoring predicts in advance heart failure admissions. International Journal of Cardiology. 2016;216:78–84. doi:10.1016/J.IJCARD.2016.04.149.

11. Seid MA, Abdela OA, Zeleke EG. Adherence to self-care recommendations and associated factors among adult heart failure patients. From the patients’ point of view. PLoS One. 2019;14(2):e0211768.

12. Ong MK, Romano PS, Edgington S, Aronow HU, Auerbach AD, Black JT, et al. Effectiveness of remote patient monitoring after discharge of hospitalized patients with heart failure: the better effectiveness after transition–heart failure (BEAT-HF) randomized clinical trial. JAMA internal medicine. 2016;176(3):310–318.

13. Van Stan JH, Mehta DD, Hillman RE. Recent innovations in voice assessment expected to impact the clinical management of voice disorders. Perspectives of the ASHA Special Interest Groups. 2017;2(3):4–13.

14. Honda K. Physiological Processes of Speech Production. Springer Handbooks. 2008; p. 7–26. doi:10.1007/978-3-540-49127-92/FIGURES/18.

15. Murton OM, Hillman RE, Mehta DD, Semigran M, Daher M, Cunningham T, et al. Acoustic speech analysis of patients with decompensated heart failure: a pilot study. The Journal of the Acoustical Society of America. 2017;142(4):EL401–EL407.

16. Center PR. Mobile Fact Sheet;. https://www.pewresearch.org/internet/fact-sheet/mobile/.

17. Portet F, Vacher M, Golanski C, Roux C, Meillon B. Design and evaluation of a smart home voice interface for the elderly: acceptability and objection aspects. Personal and Ubiquitous Computing. 2013;17(1):127–144.

18. Schlomann A, Wahl HW, Zentel P, Heyl V, Knapp L, Opfermann C, et al. Potential and pitfalls of digital voice assistants in older adults with and without intellectual disabilities: relevance of participatory design elements and ecologically valid field studies. Frontiers in Psychology. 2021; p. 2522.

19. Cleres D, Rassouli F, Brutsche M, Kowatsch T, Barata F. Lena: a Voice-Based Conversational Agent for Remote Patient Monitoring in Chronic Obstructive Pulmonary Disease. In: Joint Proceedings of the ACM IUI 2021 Workshops co-located with 26th ACM Conference on Intelligent User Interfaces (ACM IUI 2021), College Station, United States, April 13-17, 2021.. vol. 2903. sl; 2021.

20. Balasuriya SS, Sitbon L, Bayor AA, Hoogstrate M, Brereton M. Use of voice activated interfaces by people with intellectual disability. In: Proceedings of the 30th Australian Conference on Computer-Human Interaction; 2018. p. 102–112.

21. Masina F, Orso V, Pluchino P, Dainese G, Volpato S, Nelini C, et al. Investigating the accessibility of voice assistants with impaired users: mixed methods study. Journal of medical Internet research. 2020;22(9):e18431.

22. Green CP, Porter CB, Bresnahan DR, Spertus JA. Development and evaluation of the Kansas City Cardiomyopathy Questionnaire: a new health status measure for heart failure. Journal of the American College of Cardiology. 2000;35(5):1245–1255.

23. Lewis EF, Coles TM, Lewis S, Nelson LM, Barrett A, Romano CD, et al. Development, psychometric evaluation, and initial feasibility assessment of a symptom tracker for use by patients with heart failure (HFaST). Journal of patient-reported outcomes. 2019;3(1):1–15.

24. Herzstiftung S. Herzinsuffizienz;. https://swissheart.ch/erkrankungen-und-notfall/herzkrankheiten-und-hirnschlag/herzinsuffizienz.

25. Barata F, Kipfer K, Weber M, Tinschert P, Fleisch E, Kowatsch T. Towards device-agnostic mobile cough detection with convolutional neural networks. In: 2019 IEEE International Conference on Healthcare Informatics (ICHI). IEEE; 2019. p. 1–11.

26. Barata F, Tinschert P, Rassouli F, Steurer-Stey C, Fleisch E, Puhan MA, et al. Automatic recognition, segmentation, and sex assignment of nocturnal asthmatic coughs and cough epochs in smartphone audio recordings: observational field study. Journal of medical Internet research. 2020;22(7):e18082.

27. Jokić S, Cleres D, Rassouli F, Steurer-Stey C, Puhan MA, Brutsche M, et al. TripletCough: Cougher Identification and Verification From Contact-Free Smartphone-Based Audio Recordings Using Metric Learning. IEEE Journal of Biomedical and Health Informatics. 2022;26(6):2746–2757.

28. Kroenke K, Spitzer RL, Williams JB. The PHQ-9: validity of a brief depression severity measure. Journal of general internal medicine. 2001;16(9):606–613.

29. Spitzer RL, Kroenke K, Williams JB, Löwe B. A brief measure for assessing generalized anxiety disorder: the GAD-7. Archives of internal medicine. 2006;166(10):1092–1097.

30. Kroenke K, Spitzer RL, Williams JB, Löwe B. The patient health questionnaire somatic, anxiety, and depressive symptom scales: a systematic review. General hospital psychiatry. 2010;32(4):345–359.

31. Holden RJ, Karsh BT. The technology acceptance model: its past and its future in health care. Journal of biomedical informatics. 2010;43(1):159–172.

32. Ripley BD. Stochastic simulation. John Wiley & Sons; 2009.

33. Kuznetsova A, Brockhoff PB, Christensen RH. lmerTest package: tests in linear mixed effects models. Journal of statistical software. 2017;82:1–26.

34. Obuchowski NA, Lieber ML, Wians Jr FH. ROC curves in clinical chemistry: uses, misuses, and possible solutions. Clinical chemistry. 2004;50(7):1118–1125.

35. Spertus JA, Jones PG. Development and validation of a short version of the Kansas City Cardiomyopathy Questionnaire. Circulation: Cardiovascular Quality and Outcomes. 2015;8(5):469–476.

36. Liang X, Batsis JA, Zhu Y, Driesse TM, Roth RM, Kotz D, et al. Evaluating voice-assistant commands for dementia detection. Computer Speech & Language. 2022;72:101297.

37. Labfront case study: Proven expertise across a full spectrum of research;. https://www.labfront.com/case-studies.

38. Leonhard Med Secure Scientific Platform;. https://ethz.ch/staffnet/en/it-services/catalogue/server-cluster/leonhard-med-secure-scientific-platform.html.

39. Awan SN, Roy N. Toward the development of an objective index of dysphonia severity: a four-factor acoustic model. Clinical linguistics & phonetics. 2006;20(1):35–49.

40. Kane J, Drugman T, Gobl C. Improved automatic detection of creak. Computer Speech & Language. 2013;27(4):1028–1047.

41. Awan SN, Roy N, Jetté ME, Meltzner GS, Hillman RE. Quantifying dysphonia severity using a spectral/cepstral-based acoustic index: Comparisons with auditory-perceptual judgements from the CAPE-V. Clinical linguistics & phonetics. 2010;24(9):742–758.

42. Rabiner L, Schafer R. Theory and applications of digital speech processing. Prentice Hall Press; 2010.

43. Picone JW. Signal modeling techniques in speech recognition. Proceedings of the IEEE. 1993;81(9):1215–1247.

44. Bakdash JZ, Marusich LR. Repeated measures correlation. Frontiers in Psychology. 2017;8. doi:10.3389/FPSYG.2017.00456/FULL.

45. Caruana R, Niculescu-Mizil A. An empirical comparison of supervised learning algorithms. In: Proceedings of the 23rd international conference on Machine learning; 2006. p. 161–168.

46. Connor JT, Martin RD, Atlas LE. Recurrent neural networks and robust time series prediction. IEEE transactions on neural networks. 1994;5(2):240–254.

47. Graves A. Long short-term memory. Supervised sequence labelling with recurrent neural networks. 2012; p. 37–45.

48. Royston P. Multiple imputation of missing values. The Stata Journal. 2004;4(3):227–241.

49. Malarvizhi R, Thanamani AS. K-nearest neighbor in missing data imputation. International Journal of Engineering Research and Development. 2012;5(1):5–7.

50. Biessmann F, Rukat T, Schmidt P, Naidu P, Schelter S, Taptunov A, et al. DataWig: Missing Value Imputation for Tables. J Mach Learn Res. 2019;20(175):1–6.

51. WMA Declaration of Helsinki – Ethical Principles for Medical Research Involving Human Subjects – WMA – The World Medical Association;. https://www.wma.net/policies-post/wma-declaration-of-helsinki-ethical-principles-for-medical-research-involving-human-subjects/.

52. Ordinance on Human Research with the Exception of Clinical trials (HRO).;. https://fedlex.data.admin.ch/filestore/fedlex.data.admin.ch/eli/cc/2013/642/20220526/en/pdf-a/fedlex-data-admin-ch-eli-cc-2013-642-20220526-en-pdf-a.pdf.

53. Sara JDS, Maor E, Borlaug B, Lewis BR, Orbelo D, Lerman LO, et al. Non-invasive vocal biomarker is associated with pulmonary hypertension. PLoS One. 2020;15(4):e0231441.

54. Maor E, Perry D, Mevorach D, Taiblum N, Luz Y, Mazin I, et al. Vocal biomarker is associated with hospitalization and mortality among heart failure patients. Journal of the American Heart Association. 2020;9(7):e013359.

55. Amir O, Abraham WT, Azzam ZS, Berger G, Anker SD, Pinney SP, et al. Remote speech analysis in the evaluation of hospitalized patients with acute decompensated heart failure. Heart Failure. 2022;10(1):41–49.

